# Gene-environment interactions contribute to blood pressure variation across global populations

**DOI:** 10.1101/2025.07.02.25330727

**Authors:** Khushi Goda, Noah Klimkowski Arango, Francesco Tiezzi, Trudy F.C. Mackay, Fabio Morgante

## Abstract

Understanding the interplay between genetic architecture and environmental exposures is essential for elucidating the biological basis of complex traits such as blood pressure (BP). Although gene-by-environment interactions (G × E) have been previously shown to contribute to BP variation, their role in multi-ancestry cohorts remains underexplored. We hypothesize that G × E effects may explain additional variance in BP traits across diverse populations, where environmental exposures and genetic backgrounds are more heterogeneous. Here, we present an evaluation of the importance of G × E on systolic (SP), diastolic (DP), and pulse pressure (PP) in a multi-ancestry subset of 25,000 individuals from the UK Biobank. We considered 23 lifestyle variables as the environmental exposures, and estimated variance components attributed to demographics, population structure, genetic effects, environmental effects and geneby-environment interactions. Our results revealed that G × E accounts for 7% of variance in DP, 4% in SP, and 3% in PP. Notably, these estimates exceed those previously reported (2% for all BP traits) in a UK Biobank analysis restricted to White British individuals using similar lifestyle variables and methodology. However, accounting for G×E did not improve prediction accuracy in two cross-validation schemes. We also tried to uncover individual interactions affecting each trait by conducting G × E-GWAS. Although no interaction surpassed genome-wide significance, we annotated suggestive hits and uncovered genes enriched in blood pressure-relevant pathways. Our study suggests that environmental heterogeneity and diverse genetic backgrounds in multi-ancestry cohorts may amplify the role of G × E, underscoring the importance of diverse populations in capturing the full spectrum of trait architecture.

## INTRODUCTION

Understanding the genetic architecture of complex human traits is a fundamental goal of human genetics and is essential for the implementation of precision medicine. Genome-wide association studies (GWAS) have identified numerous genetic associations across a wide array of traits and diseases, yet these variants typically explain only a modest fraction of phenotypic variance^1,2^. In addition, complex traits are also affected by nongenetic factors such as diet and physical activity. For example, for blood pressure (BP) traits, the narrow sense heritability (*h*^2^) has been estimated to be 0.4-0.6^3^. However, estimates of the genomic heritability 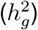 indicate that common variants explain about half of the narrow sense heritability^1^, highlighting a persistent gap, often referred to as the “missing heritability”. Finally, genetic variants identified by GWAS explain about 60% of the genomic heritability^4^. These gaps suggest the need to look beyond additive effects of common variants and consider other sources of phenotypic variation, including gene-by-environment interactions (G×E), epistasis, and rare variants. G×E, in particular, offers a biologically plausible mechanism through which environmental exposures modulate genetic risk, contributing to trait variability in ways that additive models cannot fully capture^5,6^.

Historically, quantitative genetic studies of complex traits have predominantly focused on additive genetic effects, implicitly assuming that genetic effects are the same across environments^5,6^. However, emerging evidence indicates that genetic effects can vary depending on the environmental context. G×E, where genetic effects change depending on environmental contexts, are now recognized as a critical, yet underexplored, axis in the genetic architecture of complex traits^7^. Such interactions have been extensively studied in evolutionary biology, and agricultural breeding, but only recently have large datasets become available, encompassing comprehensive genotype, phenotype, and environmental information, enabling G×E analyses for human studies. For example, environmental exposures such as dietary salt intake, alcohol use, physical activity, and sleep duration are well-established modifiers of genetic effects on BP levels^8–11^.

Despite some success, discovering G × E affecting complex traits remains challenging due to the small effect sizes of individual interactions and the substantial multiple testing burden associated with genome-wide scans^12,13^. To enhance detection power, recent studies have adopted variance component models to estimate the collective contribution of all G × E interactions to the phenotypic variance of human traits^14–19^. Unlike single-marker, single-environment approaches, which test variant-environment pairs individually, variance component models aggregate genome-wide and environment-wide signals, increasing statistical power. These models partition the total phenotypic variance 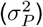 into components due to additive genetic effects 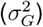, environmental effects 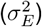, and interaction effects 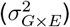 ^20–22^. Using this approach, it has been shown that G×E can explain a non-negligible proportion of phenotypic variance for several complex traits. For example, a study reported that G × E effects explained approximately 9% of variance in body mass index (BMI), 4% in systolic pressure (SP), 2% in diastolic pressure (DP), and 12% in pulse pressure (PP), highlighting the contribution of context-dependent genetic effects to variation in cardiometabolic traits^22^. Another study found evidence of gene-lifestyle interactions contributing to the variance in blood lipid levels across multiple ancestries, emphasizing the broad relevance of environmental modulation of genetic effects on metabolic traits^17^. Glycemic traits have also been shown to have substantial contributions by G × E, highlighting the role of environmental factors such as diet and physical activity in shaping genetic risk profiles for diabetes-related phenotypes^18^. Collectively, these studies illustrate the increasing recognition of context-dependent genetic effects as critical components of complex trait architecture.

The majority of genetic studies, including those investigating G × E, has predominantly focused on populations of European descent^23–25^. Genetic architectures vary across populations due to differences in allele frequencies and linkage disequilibrium (LD) patterns, which can affect the detection and interpretation of G × E signals. Specifically, variants implicated in G × E interactions may differ in their tagging efficiency across populations due to varying LD structures, resulting in certain ancestry-specific interactions remaining undetected in analyses limited to homogeneous samples^26^. Furthermore, environmental exposures differ considerably across ancestry groups due to cultural, geographic, dietary, socioeconomic, and healthcare-related factors, potentially interacting differently with genetic predispositions and exacerbating population-specific genetic mechanisms. Thus, heterogeneity in both genetic architecture and environmental exposures across ancestries may result in G × E interactions contributing more to phenotypic variance in multi-ancestry cohorts compared to single-ancestry ones^27–29^. However, this increased complexity results in statistical challenges: (1) population structure, which is exacerbated in multiancestry cohorts, can lead to biased estimates of genetic and interaction variance components if not properly accounted for^30–32^; (2) sample size imbalances across ancestries, where European-descent samples typically predominate, can reduce statistical power to detect ancestry-specific G × E interactions.

Thus, in this study, we used the UK Biobank^33^ data to quantify the contribution of G×E interactions to DP, SP, and PP across multiple ancestries, while addressing the statistical challenges posed by population structure and sample size imbalances across ancestries. We used a set of 23 lifestyle-related variables as our environmental exposures. BP traits serve as ideal model phenotypes for studying G × E across ancestries because: (1) BP is influenced by lifestyle factors such as dietary sodium intake, alcohol consumption, physical activity, sleep patterns, and socioeconomic status, making it particularly suitable for investigating context-dependent genetic effects^8–11^; (2) G×E has been shown to contribute significantly to blood pressure variation in samples of European descent^15^; (3) a multi-ancestry GWAS found low correlations of the effect sizes of significant variants across ancestries^34^; (4) the prevalence of hypertension (*i*.*e*., high blood pressure) is different across ancestries^35^.

## METHODS

### Data Extraction and Processing

Data utilized for this study were sourced from the UK Biobank (UKBB) resource, a large-scale biomedical database that contains extensive genotype and phenotype data. All analyses and data processing steps were performed using R v4.2.3 and Python v3.9.5.

#### Phenotype, Lifestyle, and Demographic Data

The UK Biobank includes phenotypic data from approximately 500,000 participants residing in the United Kingdom^33^. Participants from multiple ancestry backgrounds were included and grouped into six broad self-identified ancestries: (1) White British, White Irish or Other White (White, *n*_*w*_ = 472, 656); (2) Mixed ethnicity including White and Black Caribbean or White and Black African or White and Asian or Other Mixed (Mixed, *n*_*m*_ = 2, 956); (3) Asian or Asian British including Indian or Pakistani or Bangladeshi or Other Asian (Asian, *n*_*a*_ = 9, 880); (4) Black or Black British, including Caribbean or African or Other Black (Black, *n*_*b*_ = 8, 060); (5) Chinese (Chinese, *n*_*c*_ = 1, 573); (6) Other ethnic groups (Other, *n*_*o*_ = 4, 558). To facilitate interpretation, we excluded individuals in the Other ethnic group for our study.

The BP phenotypes analyzed included the automated readings of DP and SP at the first assessment. Individuals undergoing medication treatment for BP had their systolic and diastolic measurements adjusted to account for medication effects. Specifically, for individuals on anti-hypertensive medications, 15 mmHg was added to their SP measurements and 10 mmHg was added to their DP measurements^15,36^. We computed PP as the difference between adjusted SP and DP.

Demographic variables (*d* = 3) included self-identified sex, age at phenotyping, and age^2^. Only individuals whose age at the time of phenotyping fell within the range of 40 to 70 years were included in our analysis, as in previous studies^15^. A set of *l* = 23 lifestyle variables was selected based on their established relationships with BP traits^15,37^, with additional consideration given to reducing multicollinearity and maximizing sample size across multiple ancestries (given the large amount of missing values). The selected variables were: Townsend deprivation index (Townsend), moderate physical activity days per week (act0 d), television watching duration (TVtime), sleep duration (sleep d), deviation in sleep duration (sleep dev; computed as the squared deviation from the mean sleep duration across individuals), current smoking status (smoking now), cooked vegetable intake (veg cook), oily fish intake (fish oily), non-oily fish in-take (fish lean), processed meat intake (meat proc), poultry intake (poultry), beef intake (beef), lamb intake (lamb), pork intake (pork), cheese intake (cheese), salt intake (salt), tea consumption (tea), alcohol frequency (alc1), waist circumference (waist), ease of getting up (getup), coffee consumption (coffee), past smoking status (smoked past), body fat percentage (BFP).

Individuals without complete records across phenotypic, demographic, or lifestyle information were excluded from the analyses. All variables underwent preprocessing steps to remove outliers, following previous studies^15,38^. Individuals with phenotypic values deviating beyond four standard deviations from the mean were removed. Individuals with values exceeding the 99th percentile for coffee, tea, veg cook, TVtime, and sleep d or below the 1st percentile for sleep d were removed. After filtering, we were left with *n*_*w*_ = 309, 932 White individuals, *n*_*m*_ = 1, 746 Mixed individuals, *n*_*a*_ = 4, 957 Asian individuals, *n*_*b*_ = 3, 979 Black individuals, *n*_*c*_ = 774 Chinese individuals. Phenotypes (DP, SP, PP) were scaled to mean 100 and standard deviation 10, while lifestyle variables and demographic variables were centered and scaled to have mean 0 and standard deviation 1.

#### Genotype Data

Genotype array data were initially processed separately for each ancestry group, including Black, Asian, Mixed, Chinese, and White populations, using PLINK v1.9^39^. We retained genetic variants meeting the following criteria in each ancestry – minor allele frequency (MAF) greater than or equal to 0.01, genotype missingness rate (GENO) less than 0.1, Hardy-Weinberg equilibrium (HWE) p-value greater than 10^*−*10^, and minor allele count (MAC) greater than or equal to 5. This filtering procedure resulted in *p* = 302, 688 autosomal genetic variants for the following analyses. To mitigate confounding from genetic relatedness, we pruned individuals based on pairwise relationships using PLINK v1.9, such that no pairs of individuals within each ancestry group had relationship coefficient greater than 0.05. This procedure resulted in *n*_*w*_ = 253, 459 White individuals, *n*_*b*_ = 3, 347 Black individuals, *n*_*a*_ = 4, 120 Asian individuals, *n*_*m*_ = 1, 265 Mixed individuals, and *n*_*c*_ = 746 Chinese individuals. To avoid disproportionate representation of White individuals and maintain a balanced multi-ancestry dataset, we randomly sampled *n*_*w*_ = 15, 000 White individuals from their total. The final total sample size for subsequent analyses was *n* = 24, 478. The final sample sizes across ancestry groups are summarized in Figure 1D.

**Figure 1.**
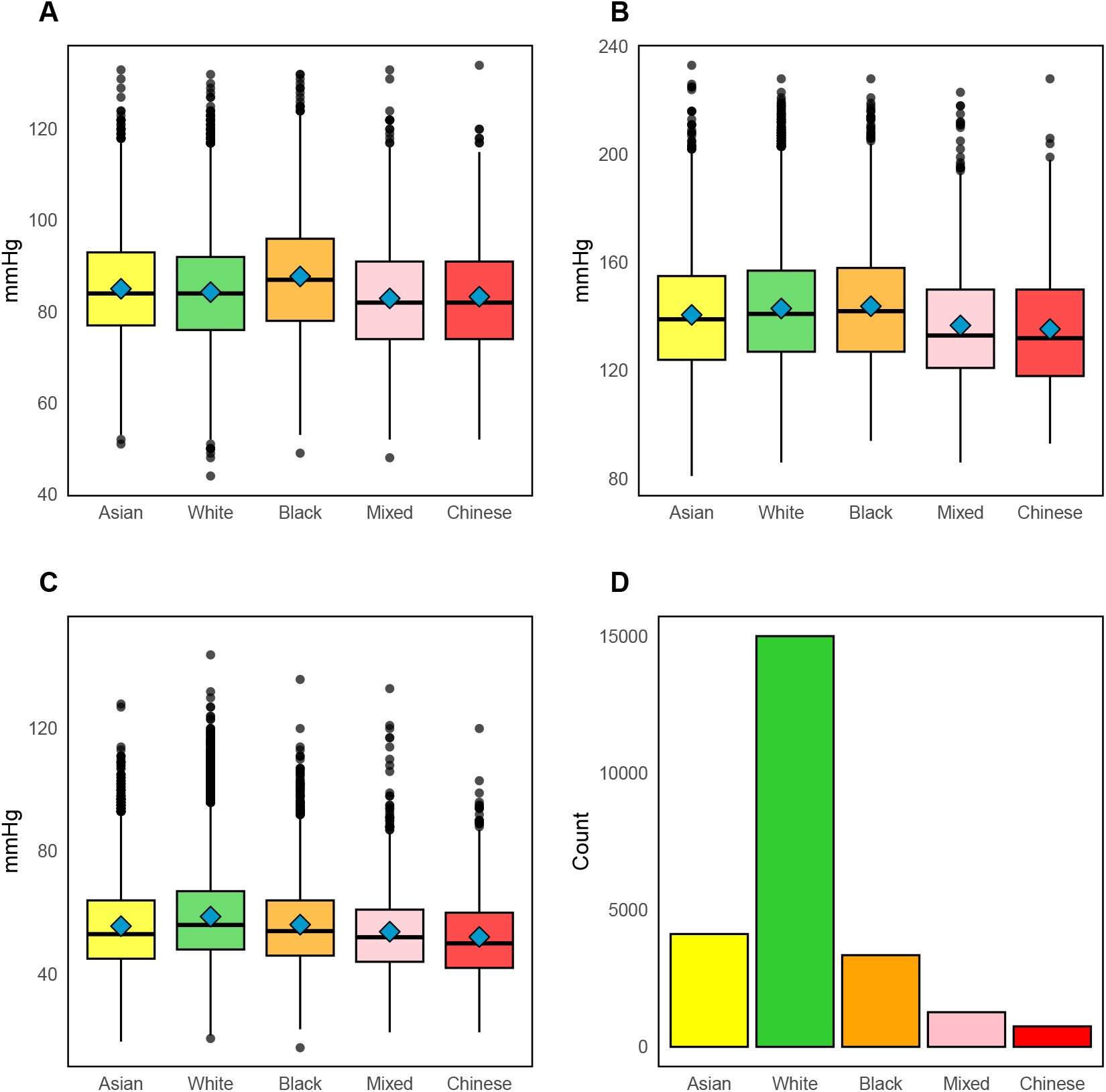
Distribution of blood pressure traits by ancestry. **(A)** Diastolic pressure (DP), **(B)** Systolic pressure (SP), and **(C)** Pulse pressure (PP) after medication adjustment. **(D)** Final sample size across self-reported ancestry groups after all filtering and sampling steps.

### Statistical Analysis

#### Population Structure

To investigate genetic population structure in our sample, we conducted Principal Component Analysis (PCA) of the genotype data^40,41^. In particular, we computed the genomic relationship matrix (GRM) as in^42^ using PLINK v1.9 and performed its eigen decomposition. To investigate environmental population structure, we performed PCA of the lifestyle data, by doing the eigen decomposition of the environmental kernel computed as in^21^ and described in the next section.

#### Estimation of Variance Components

We estimated the proportion of BP variance explained by different components (*i*.*e*., demographic factors, population structure, genetic predisposition, lifestyle factors, gene-lifestyle interactions) using the Reproducing Kernel Hilbert Spaces (RKHS) regression approach^43^. We used the implementation of this framework available in the BGLR v1.1.2 R package, which adopts a Bayesian approach to the estimation of all the model parameters^44^.

We implemented six models:

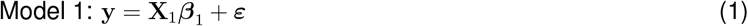

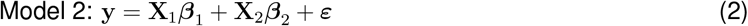

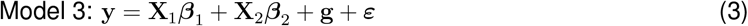

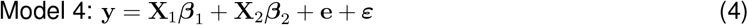

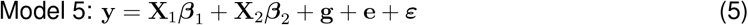

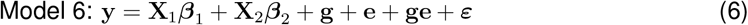

where,

**y** is a *n*-vector of phenotypic values,

**X**_1_ is a *n* ×*d* design matrix of demographic variables (sex, age, age^2^), ***β***_1_ is a *d*-vector of their “fixed” effects,

**X**_2_ is a *n* ×*c* design matrix including the top *c* = 10 genetic principal components, ***β***_2_ is a *c*-vector of their “fixed” effects,

**g** is a *n*-vector of random additive genetic values, 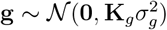,

**e** is a *n*-vector of random lifestyle values, 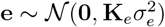,

**ge** is a *n*-vector of random interaction values, 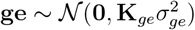,

***ε*** is a *n*-vector of random residual values,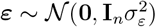.

The kernel **K**_*g*_ is *n × n* matrix, where each off-diagonal element denotes the coefficient of genetic relationship between each pair of individuals. This is also known as the GRM. The standard approach to compute the GRM^42,45^ captures both recent genetic relatedness and distant population structure, when the latter is present such as in the present study^31^. Since we were interested in estimating the variance explained by genetic variants without the contribution of population structure, we computed **K**_*g*_ using the Principal Components-Adjusted Relatedness Estimation (PC-Relate) approach implemented in the GENESIS v2.24.2 R package. This approach removes the contribution of distant population structure from estimates of genetic relationships using the top genetic principal components (PCs)^31^. In this study, we used the top 5 PCs computed using the PC-AiR method^46^. The kernel **K**_*e*_ is a *n*× *n* matrix, where each off-diagonal element denotes the similarity of each pair of individuals based on the lifestyle variables. It was computed as **EE**^⊺^*/l*, where **E** is a *n* × *l* matrix of lifestyle variables^21^. The kernel **K**_*ge*_ is a *n × n* matrix computed as **K**_*g*_ ∘**K**_*e*_, where ∘denotes the Hadamard product. This kernel captures gene-lifestyle interactions^21^.

The “fixed” effects were assigned uniform priors (which results in unshrunken estimates) and the variance components were assigned inverted *χ*^2^ priors^44^. The models were run for a total of 90,000 iterations, discarding the initial 40,000 iterations as burn-in, followed by thinning every 50 iterations. Convergence was established by visual inspection of the trace plots of all model parameters. Variance components – computed as described in^37^ – were summarized with the posterior mean as a point estimate and the 95% empirical credible interval as a measure of error.

#### Identification of individual interactions via G×E-GWAS

To identify genetic variants whose effects on BP were modulated by lifestyle variables, we performed variant-by-lifestyle genome-wide association studies (G×E-GWAS). These analyses were conducted using PLINK v2.047 <monospace>--glm interaction <monospace>argument. For each genetic variant and each lifestyle variable, we fit the following model:

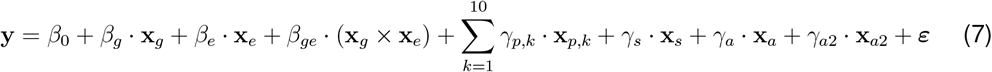

where,

**y** denotes the BP phenotype (DP, SP or PP),

**x**_*g*_ is *n*-vector of genotypes,

**x**_*g*_ is *n*-vector of lifestyle variable values,

**x**_*pk*_ is *n*-vector of values for the *k*^*th*^ principal component,

**x**_*s*_ is *n*-vector of sex indicators,

**x**_*a*_ is *n*-vector of age values,

**x**_*a*2_ is *n*-vector of age^2^ values,

*β*_*g*_, *β*_*e*_, *β*_*ge*_, *γ*_*p,k*_, *γ*_*s*_, *γ*_*a*_, *γ*_*a*2_ are the respective effects,

*β*_0_ is the intercept.

The interaction effect *β*_*ge*_ was evaluated for significance, and p-values associated with this term were used to identify genetic variants exhibiting significant lifestyle variable-dependent effects. Significance was evaluated by inspecting Quantile-Quantile (QQ) plots for each lifetyle variable, identifying exposures where observed p-values exhibited notable deviation from the expected null distribution for small p-values. For lifestyle variables displaying substantial inflation, we prioritized genetic variants with the lowest p-values based on their departure from the expectation of no association. These top genetic variants were annotated using the GWAS Catalog^48^. Variants were then annotated using Ensembl Variant Effect Predictor (VEP)^49^ on the GRCh37 (hg19) reference genome build^50^ to identify genes of interest. To further characterize biological relevance, we conducted functional enrichment analyses using g:Profiler^51^ to systematically evaluate whether genes identified through suggestive G ×E signals converged on shared biological processes or pathways. Functional enrichment was performed against the full annotated human genome as the statistical background. Significance was assessed using the g:SCS (Set Counts and Sizes) threshold (*α <* 0.05), the default multiple testing correction in g:Profiler. Only terms that passed the g:SCS threshold were considered significant^52^. We further crossreferenced annotated genes against BP, cardiovascular and hypertension trait associated loci curated in the GWAS Catalog^53^.

#### Prediction of Blood Pressure Traits

We evaluated the predictive ability of the models using two cross-validation (CV) approaches: random CV and across-ancestry CV. In the random CV, the individuals were randomly partitioned into five equal subsets (folds). Iteratively, each fold served once as the validation set, while the remaining four folds formed the training set. In the across-ancestry CV, individuals were grouped according to ancestry – White, Black, Asian, Mixed, and Chinese. Iteratively, data from all but one ancestry formed the training set, and the omitted ancestry group formed the validation set. We fitted the same models described above to the training set and used the estimated parameters to predict the phenotypes in the validation set. Prediction accuracy was computed as the *R*^2^ from the regression of the true phenotypes on the predicted phenotypes of the validation set individuals.

## RESULTS

### Distribution of Blood Pressure Traits Across Ancestries

Figure 1 A–C shows the distribution of three BP phenotypes— DP, SP, and PP—across ancestry groups.

DP (Figure 1A) showed variability across groups, with Black individuals exhibiting the highest mean DP (87.4 mmHg) and Chinese individuals the lowest (82.9 mmHg). SP (Figure 1B) showed similar differences across ancestries, with means ranging from 135.4 mmHg in Chinese to 143.8 mmHg in Black individuals. PP (Figure 1C) followed a different pattern, with White individuals showing the highest mean PP (58.7 mmHg) and Chinese individuals the lowest (52.1 mmHg). These results highlight the presence of phenotypic variability across ancestries for the three BP traits.

### Population Structure

The first two genetic principal components (PC1 and PC2), visualized in Figure 2A, showed that there was substantial population structure in our sample, as expected given the presence of individuals from different self-reported ancestries.

**Figure 2.**
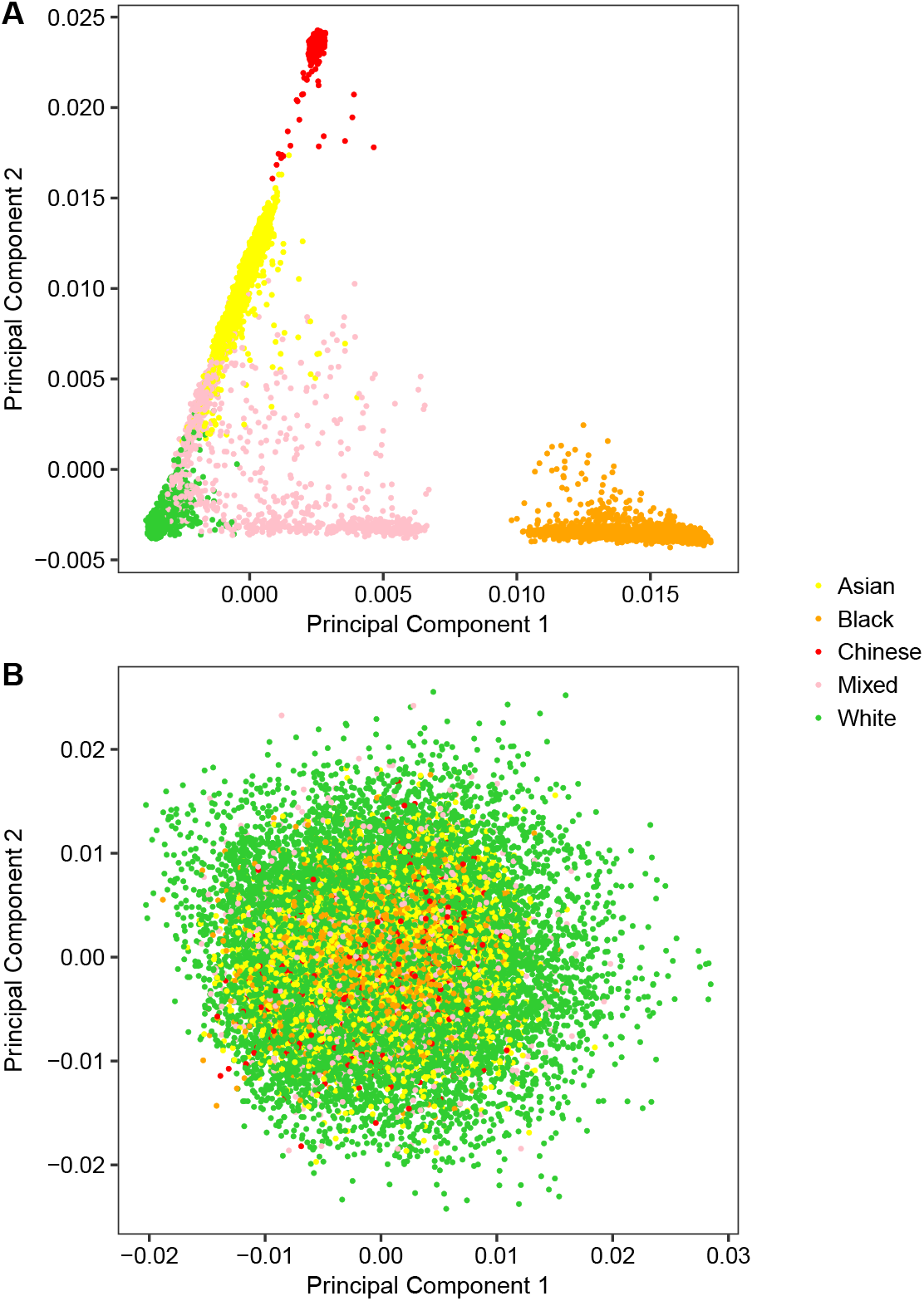
Principal Component Analysis of the genotype data based on (A) the commonly used GRM and (B) the PC-Relate GRM. Individuals are colored by self-reported ancestry.

The presence of population structure is a problem when trying to estimate genetic variance components modeled using the GRM computed as in^42^. This GRM captures both recent genetic relatedness and distant population structure, producing estimates of the genetic variance components that are inflated by structure^31,54^. To overcome this, we constructed a GRM using the PC-Relate framework, which estimates genetic relationships corrected for population structure^31^. PCA of the PC-Relate-based GRM (Figure 2B) shows a more homogeneous distribution in PC space, with no clear separation among ancestries. Thus, we used the PC-Relate-based GRM in all the subsequent analyses.

On the other hand, the lifestyle data did not show patterns of population structure as strong as the genetic data, although some differences among ancestries seemed to exist (Figure 3).

**Figure 3.**
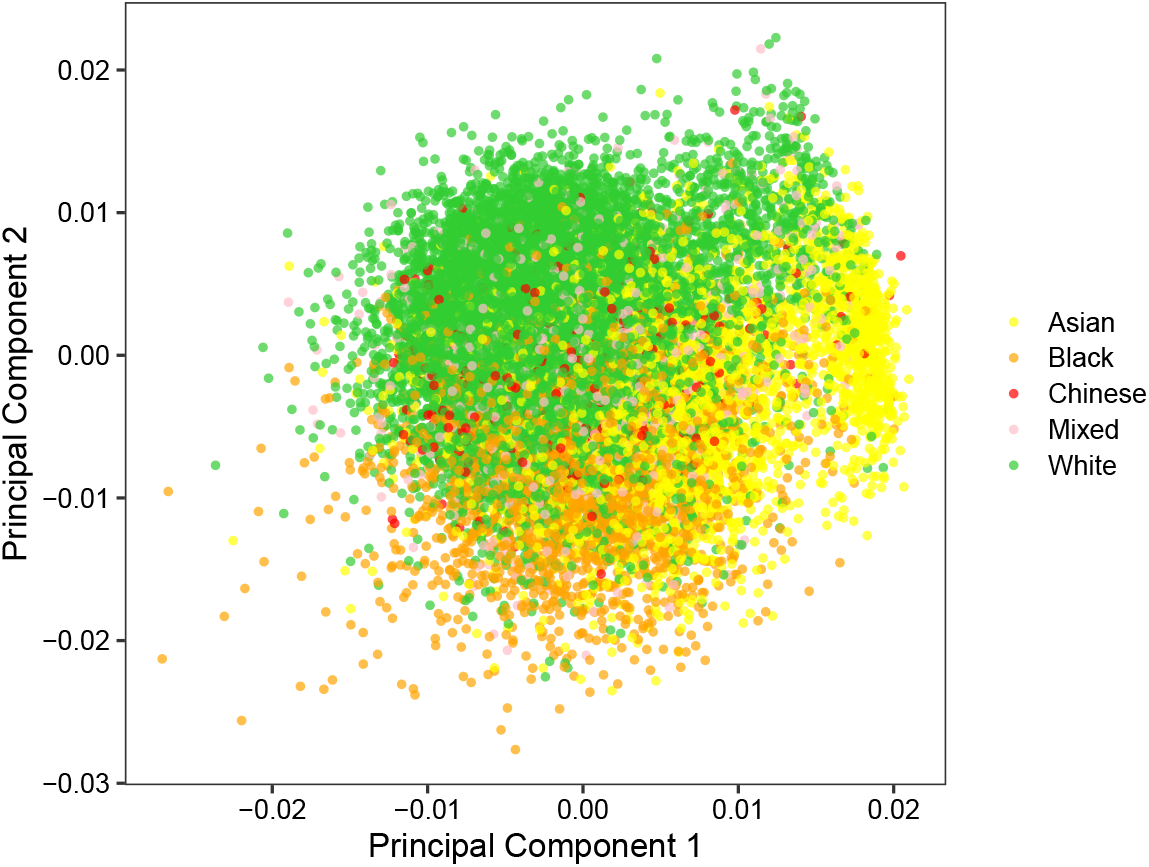
Principal Component Analysis of the lifestyle data. Individuals are colored by self-reported ancestry.

### Estimation of Variance Components

Figure 4 presents the estimated variance components across the six models used.

**Figure 4.**
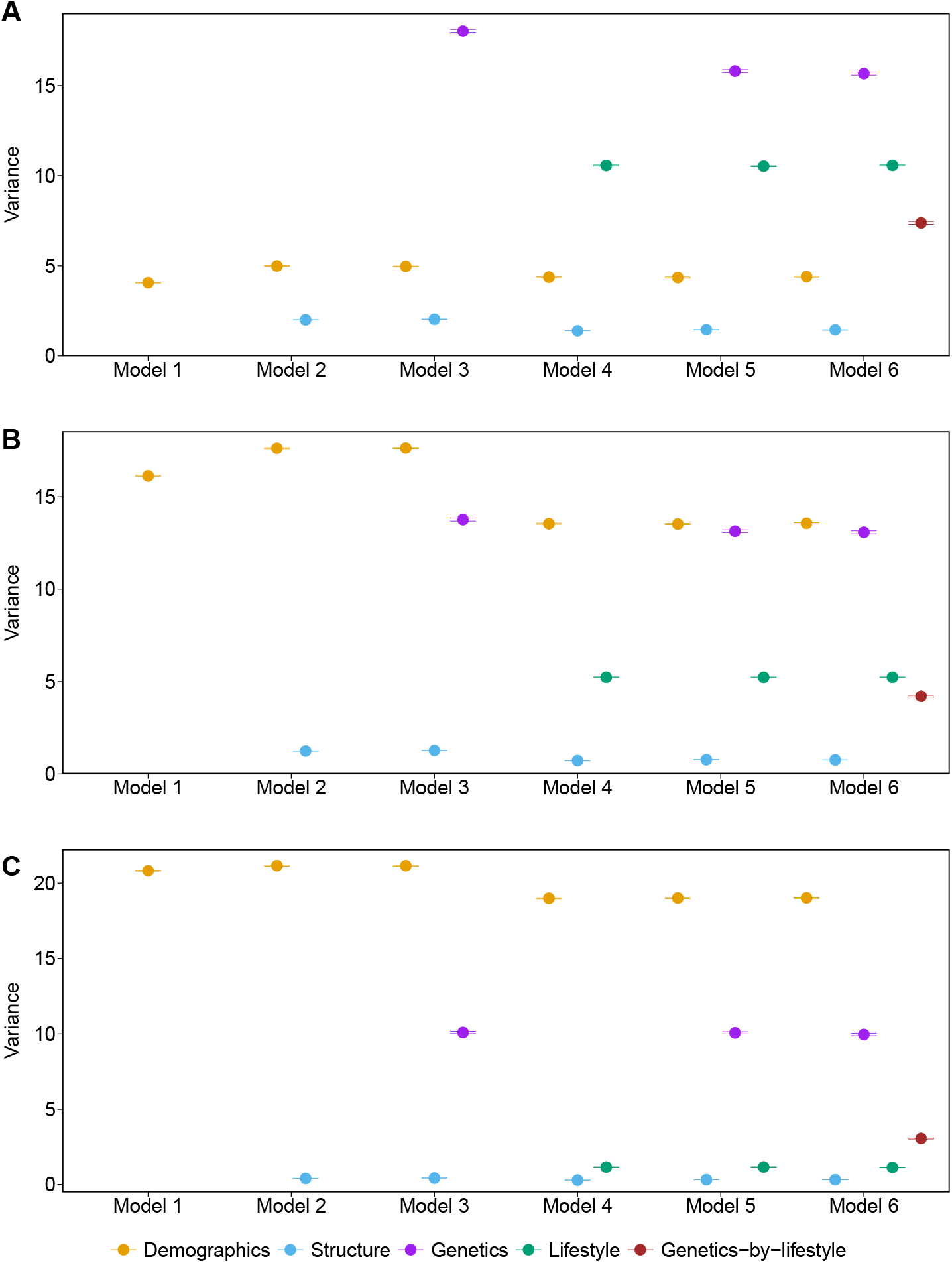
Estimates of the variance components with six models for the blood pressure traits. **(A)** Diastolic pressure (DP), **(B)** Systolic pressure (SP), and **(C)** Pulse pressure (PP). Bars represent the 95% empirical credible interval for the posterior distribution.

Model 1, which included only demographic variables (sex, age, and age^2^), explained 4.05% of the variance in DP, 16.13% in SP, and 20.83% in PP. These values represent the baseline contribution of demographic factors to blood pressure traits variation. Incorporating genetic principal components in Model 2 increased the explained variance to 7.00%, 18.87%, and 21.57% for DP, SP, and PP, respectively. This increase is attributed to the top 10 PCs, which marginally explained 2.00% of DP variance, 1.24% of SP variance, and 0.40% of PP variance, underscoring the limited influence of population structure to these traits. Model 3 introduced the additive genetic effects, leading to a substantial increase in total variance explained: 25.03% for DP, 32.67% for SP, and 31.68% for PP. The genetic effect alone contributed 18.02% of the DP variance, 13.76% of the SP variance, and 10.09% of the PP variance. Model 4, which instead incorporated lifestyle effects in addition to demographic variables and genetic PCs, explained 16.32% of variance in DP, 19.49% in SP, and 20.44% in PP. Of this, the lifestyle effect contributed 10.56% of DP variance, 5.24% of SP variance, and 1.16% of PP variance. Model 5 included both genetic and lifestyle effects as well as demographic variables and genetic PCs, increasing the total variance explained to 32.12% for DP, while slightly reducing it to 32.65% for SP and 30.56% for PP compared to Models 3 and 4. The genetic component explained 15.80% of DP variance, 13.13% of SP variance, and 10.07% of PP variance, while the lifestyle component contributed 10.52%, 5.23%, and 1.16% of DP variance, SP variance, and PP variance, respectively. Model 6 additionally incorporated the gene-lifestyle interactions, resulting in the highest overall variance explained across all traits: 39.45% for DP, 36.83% for SP, and 33.49% for PP. The G × E component alone accounted for 7.37%, 4.20%, and 3.05% of the variance in DP, SP, and PP, respectively. These findings support the hypothesis that G × E explain a substantial portion of phenotypic variance in BP.

Variance component estimates remained stable across the models in which they were included, with the exception of the demographic component for SP and, to a lesser extent, PP. In fact, the estimate of the variance explained by demographics diminished in the models that also included the lifestyle component (from ∼16*/*17% for SP and ∼21% for PP with Model 1-3 to ∼13% for SP and ∼19% for PP with Model 4-6). The estimate of the genetic variance component for DP also got slightly reduced (from 18.02% to 15.60%) when including lifestyle information as well (Model 3 vs Model 5,6). This phenomenon may be due to the presence of correlations between demographic and lifestyle variables, and genetic and lifestyle variables, respectively.

### Identification of individual interactions via G×E-GWAS

To identify specific loci exhibiting evidence of gene-by-lifestyle interaction effects on BP traits, we conducted genome-wide interaction testing between 302,688 genetic variants and 23 lifestyle variables across three BP traits. For each lifestyle–trait combination, we defined genome-wide significance using a Bonferroni correction threshold of *p <* 7.18 ×10^*−*9^, correcting for the total number of tests, *i*.*e*., 302, 688 ×23. All lifestyle–trait combinations yielded no significant genetic variants with this threshold, except for TVtime. Six genetic variants passed the threshold for SP and one genetic variant for PP. However, visual inspection of the QQ plots for SP and PP revealed evidence of widespread inflation across the p-value distribution, deviating from the null expectation. This is consistent with uncorrected confounding. As such, we excluded TVtime from downstream annotation and enrichment analyses.

To further prioritize G × E associations that did not meet a Bonferroni-corrected significance threshold, but exhibited signs of potential true signal, we visually inspected QQ plots for each lifestyle–trait combination. Inflation beyond the expectation under the null hypothesis of no association for small p-values was used to identify lifestyle variables likely to harbor suggestive G × E signals. If QQ plots showed strong inflation at large p-values, which is a sign of uncorrected confounding, we did not consider those lifestyle variables. For lifestyle variables whose QQ plots exhibited an evident change of slope (“elbow”) near the smallest p-values (even if there was moderate inflation at larger p-values), we selected a conservative p-value cutoff corresponding to the onset of this deviation—typically in the range of 10^*−*4^ to 10^*−*6^—to retain a subset of top-ranked variants for downstream annotation and gene prioritization. This included lifestyle variables BFP, act0 d, cheese, poultry, and pork for DP (Figure S1). For SP (Figure S2), coffee, act0 d, veg cook, tea, and Townsend were selected. For PP (Figure S3), only waist, Townsend, and sleep d were selected. Table 1 outlines the selected lifestyle variables for each trait and the number of genetic variants included in downstream functional annotation.

**Table 1.**
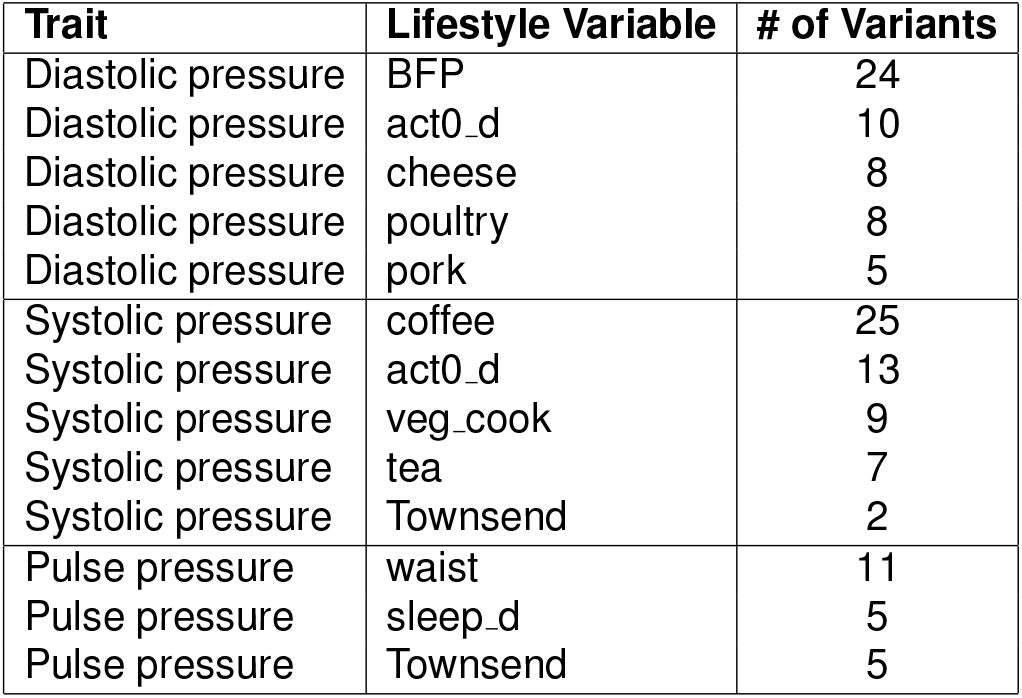
Number of genetic variants selected for downstream annotation by trait and lifestyle variable.

| Trait | Lifestyle Variable | # of Variants |
| --- | --- | --- |
| Diastolic pressure | BFP | 24 |
| Diastolic pressure | act0_d | 10 |
| Diastolic pressure | cheese | 8 |
| Diastolic pressure | poultry | 8 |
| Diastolic pressure | pork | 5 |
| Systolic pressure | coffee | 25 |
| Systolic pressure | act0_d | 13 |
| Systolic pressure | veg_cook | 9 |
| Systolic pressure | tea | 7 |
| Systolic pressure | Townsend | 2 |
| Pulse pressure | waist | 11 |
| Pulse pressure | sleep_d | 5 |
| Pulse pressure | Townsend | 5 |

The complete lists of variants (Table S1 and Table S2), genes (Table S3), and GO terms (Table S4) for all traits are available in the supplement. Here, we highlight a few interesting examples.

For DP, the selected variants included known associations with lipid levels and body composition. For example, rs4781668 (cheese) in BMERB1 has known associations with cholesterol ester levels, phosphatidylcholine levels, and height^55,56^. rs6719772 (pork) in LRRTM4 is connected to lipid levels in elite athletes^57^. rs4675569 (act0 d) in PPIAP68 is associated with striated muscle fiber area^58^. VEP analysis identified 7 genes related to the selected variants. GWAS Catalog associations for these genes include vascular biology, blood metabolites, immune system function, and neurological disorders. For example, KCNMB3, a calcium-activated potassium channel component, has known associations with corpuscular hemoglobin, antidepressant response, and electrocardiogram (EKG) responses to Trypanosoma infection^59–61^. TLR10, a tolllike receptor, has known associations with peripheral artery disease and asthma as part of the immune system^62,63^. AP2A2, an Adaptor protein complex subunit, is associated with cadherin levels, white blood cell response, and Alzheimer’s Disease^64–66^. Functional enrichment analysis identified the CORUM term PNP homotrimer complex (CORUM:6208). Further exploration of the CORUM term reveals two GO terms related to purine nucleoside metabolism (GO:0004731, GO:0042278) and one term for regulation of T cell proliferation (GO:0042129).

For SP, selected variants included known associations for vascular biology, blood metabolites, and acute myeloid leukemia. rs13053817 (coffee) in RFPL1S (Ret finger protein like antisense) has been associated with carotid artery thickness^67^ and rs4520040 (veg cook) in R3HDM2P2 (R3H domain pseudogene) with cardiac hypertrophy^68^. rs786906 (veg cook) in PKN2, a protein kinase, has known associations to systolic pressure and intraocular pressure^69,70^. Additionally, rs12145922 (veg cook) in PKN2-AS1, an antisense RNA to PKN2, has known associations to red blood cell counts, liver enzyme levels, and sphingomyelin levels^57,71,72^. VEP analysis identified 43 genes related to the selected variants. These genes are involved in core blood pressure phenotypes as well as immunological proteins and disorders, heart disease, neurological disorders, and the gut microbiome. SCARB1, a lipid scavenger receptor, is an established gene related to high density lipoprotein levels and coronary artery disease^64,73^. TLR10, a toll-like receptor, has known associations to peripheral artery disease and asthma as part of the immune system^62,63^. TXNDC16, a thioredoxin domain containing protein, is primarily involved in bromotryptophan levels^74^, as well as tinnitus, Alzheimer’s Disease, arthritis, sleep loss, and memory loss^66,75–78^. PPM1H, a magnesium dependent protein phosphatase, is associated with gut microbiome characteristics, red blood cell physiology, obsessive-compulsive disorder, and heart failure^72,79–81^. Functional enrichment analysis discovered 8 GO terms related to the clathrin coating process for the Golgi apparatus.

For PP, none of the selected variants had known associations in the GWAS Catalog. VEP identified 6 genes related to neurological disorders, core blood pressure phenotypes, and blood metabolite levels. DAB1, a neuronal growth regulator, has several associations to phenotypes including restless leg syndrome, smoking initiation, insomnia, antihypertensive medication use, and tumor calcium signaling^64,72,82,83^. SMYD1, a muscle-specific histone deacteylase, has associations for alanine levels, night sleep phenotypes, neurofibrillary tangles, retinal detachment, and vascular growth factor levels^84–88^. Functional enrichment analysis identified one GO term for voltage-gated calcium channel activity (GO:0086057) and one CORUM term for the LRRC8ALRRC8D complex (CORUM:6582). The CORUM term is associated with two GO terms: voltagegated ion channel activity (GO:0005225) and cell volume homeostasis (GO:0006884).

### Prediction of Blood Pressure Traits

We then sought to use the models described above to predict yet-to-be-observed phenotypes. To evaluate the generalizability and transferability of the models across genetically diverse populations, we conducted a cross-ancestry prediction analysis. In this setting, models were trained on all but one ancestry group and then tested on the held-out group. Figure 5 presents *R*^2^ values across the six models (Model 1 through Model 6) for each BP trait and ancestry group.

**Figure 5.**
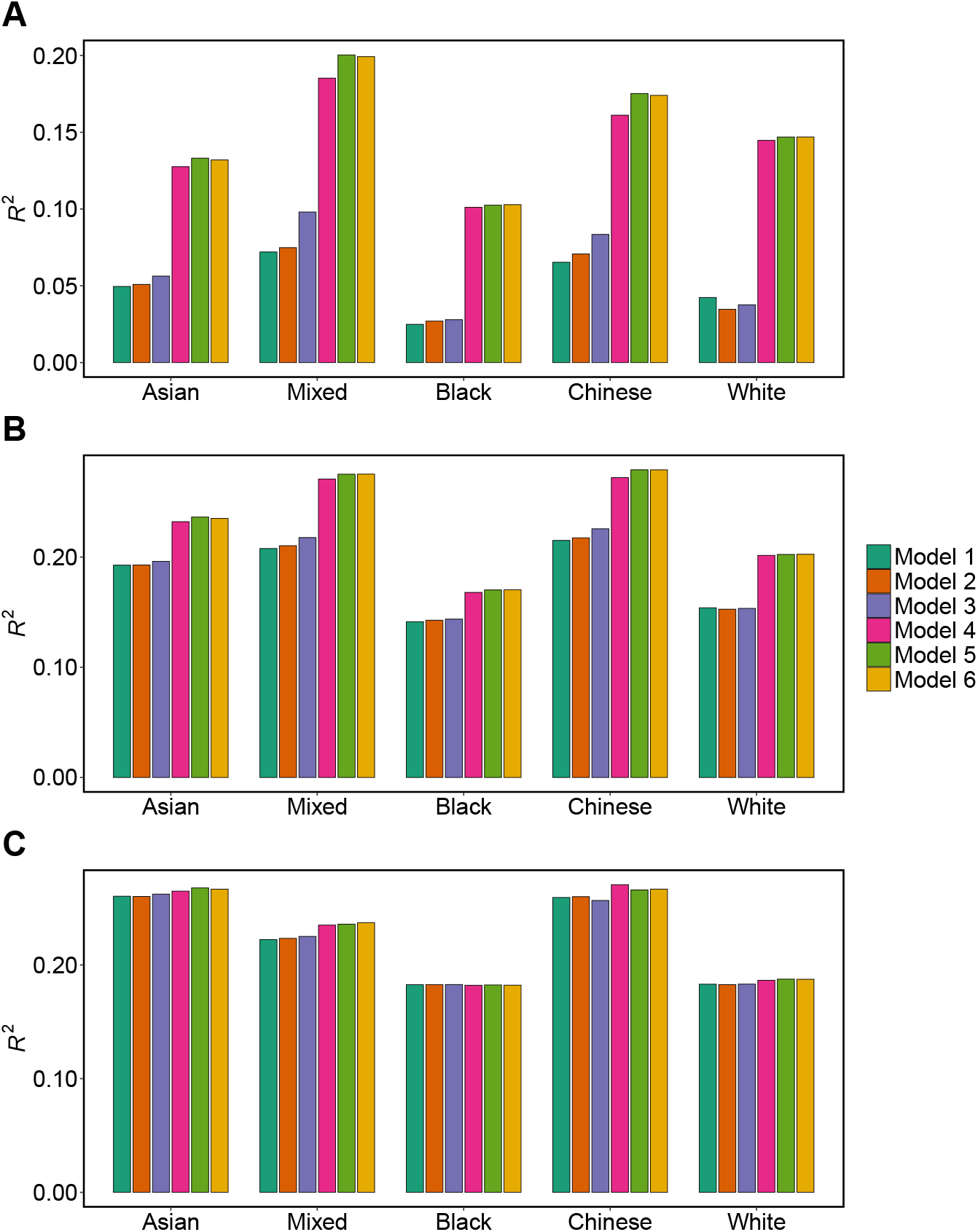
Cross-ancestry prediction performance of blood pressure traits. *R*^2^ values shown for six predictive models (Model 1 to Model 6) trained on individuals from all but one ancestry group and tested on the held-out group. The group in the validation set is denoted on the x-axis. **(A)** Diastolic pressure (DP), **(B)** Systolic pressure (SP), and **(C)** Pulse pressure (PP).

Prediction accuracy did not improve uniformly across all models and traits. Trait-specific patterns emerged for Model 1. DP exhibited lowest baseline predictability, followed by SP and then PP, which showed the highest prediction accuracy, in line with the proportion of variance explained by demographic variables. For example, for Asian ancestry, prediction accuracy was *R*^2^ = 0.260 for PP and *R*^2^ = 0.193 for SP, but only *R*^2^ = 0.0.050 for DP. *R*^2^ for Model 1 was highest in PP for Asian (*R*^2^ = 0.260) and Chinese (*R*^2^ = 0.259) ancestry, followed by Mixed (*R*^2^ = 0.222) ancestry. Black and White ancestries showed the lowest *R*^2^ of 0.183 for Model 1. Incorporating population structure via principal components (Model 2) yielded negligible gains over the baseline demographic model (Model 1) across all traits and ancestries. Model 3, which introduced additive genetic effects, provided an improvement in prediction accuracy for DP in individuals of Asian (*R*^2^ = 0.056), Mixed (*R*^2^ = 0.098), and Chinese (*R*^2^ = 0.083) ancestries, and for SP in Mixed (*R*^2^ = 0.218) and Chinese (*R*^2^ = 0.226) ancestries. However, no substantial gain from Model 3 was observed for PP across any ancestry group. DP exhibited the most pronounced improvement from the inclusion of lifestyle effects (Model 4). For example, in Asian, predictive accuracy increased from *R*^2^ = 0.050 in Model 1 to *R*^2^ = 0.128 in Model 4; in Black, from *R*^2^ = 0.025 to *R*^2^ = 0.101; and in Mixed group, from *R*^2^ = 0.072 to *R*^2^ = 0.185. SP followed a similar trend. In Chinese, *R*^2^ increased from 0.215 in Model 1 to 0.272 in Model 4, and in White from *R*^2^ = 0.154 to *R*^2^ = 0.202. PP displayed similar predictive performance across models, with modest gains from adding additional effects beyond that of demographics, consistent with a large proportion of variance explained by demographic variables. For example, *R*^2^ improved from 0.260 to 0.271 in Chinese and from 0.222 to 0.235 in Mixed from Model 1 to Model 4.

For DP, the highest prediction accuracy was observed when genetic and lifestyle components were jointly modeled in Model 5, emphasizing their (at least partly) orthogonal contribution to predictions. *R*^2^ was equal to 0.103 in Black, 0.147 in White, 0.133 in Asian, 0.200 in Mixed, and 0.175 in Chinese ancestries. For SP, the performance of Model 5 was much closer to that of Model 4, with *R*^2^ values only marginally higher in Model 5 (*R*^2^ = 0.170 in Black, *R*^2^ = 0.202 in White, *R*^2^ = 0.236 in Asian, *R*^2^ = 0.275 in Mixed, and *R*^2^ = 0.279 in Chinese ancestries). For PP, prediction accuracy from Model 5 was marginally higher than Model 4 only in Asian (*R*^2^ = 0.268). For the other ancestries, prediction accuracy from Model 5 was the same or even lower (*R*^2^ = 0.266 in Chinese) than Model 4. By contrast, the inclusion of G × E in Model 6 resulted in no additional improvement for all the trait/ancestry combinations.

Comparing prediction accuracies across ancestries, our results showed systematic differences. In fact, the Black ancestry consistently exhibited the lowest *R*^2^ values, followed by White and Asian ancestries. In contrast, Mixed (except for PP) and Chinese achieved the highest predictive performance.

In addition to cross-ancestry predictions, we conducted 5-fold cross-validation by randomly assigning individuals to folds irrespective of ancestry. Figure 6 presents *R*^2^ values across six hierarchical models (Model 1 through Model 6) for each BP trait.

**Figure 6.**
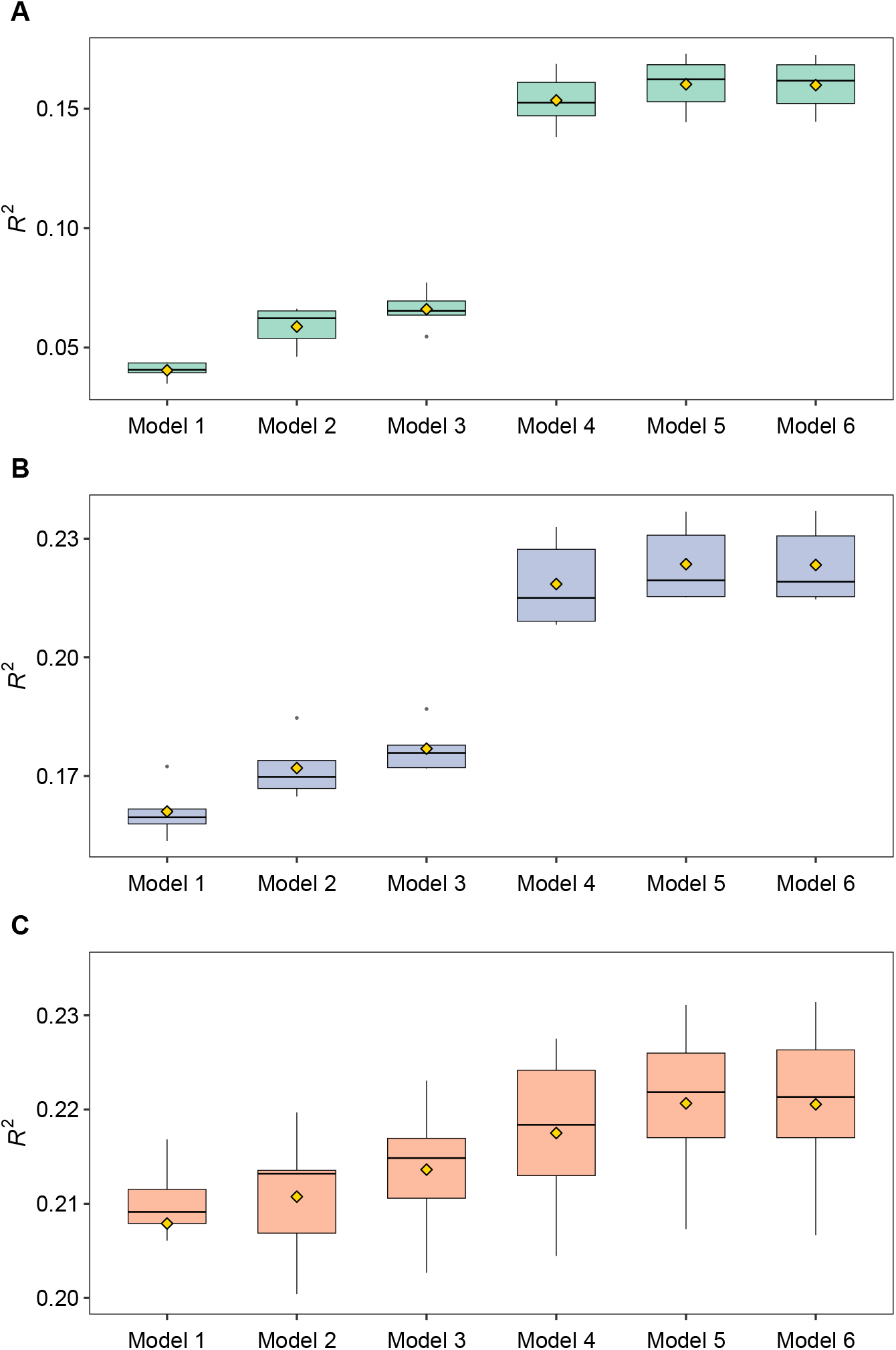
Random cross-validation prediction performance for blood pressure traits. Predictive accuracy (*R*^2^) across five random cross-validation folds for each of six models (Model 1 to Model 6). **(A)** Diastolic pressure (DP), **(B)** Systolic pressure (SP), and **(C)** Pulse pressure (PP).

For DP, the trait with the lowest impact from demographic variables, predictive accuracy improved across models from *R*^2^ = 0.040 in Model 1 to *R*^2^ = 0.059 in Model 2, and further to *R*^2^ = 0.066 in Model 3. Model 4 increased performance substantially to *R*^2^ = 0.153, while Model 5 slightly increased it to *R*^2^ = 0.160. Including G × E interaction terms in Model 6 did not improve accuracy further. For SP, a trait with higher impact from demographic variables and lower influence from lifestyle compared to DP, *R*^2^ improved from 0.161 in Model 1 to 0.172 in Model 2, and to

0.177 in Model 3. Model 4 improved prediction accuracy substantially, providing *R*^2^ = 0.219, while Model 5 improved accuracy only marginally (*R*^2^ = 0.224). Again, Model 6 had virtually identical accuracy to Model 5. On the other hand, PP showed the highest baseline predictive performance from demographic variables (*R*^2^ = 0.208 in Model 1) among the three traits, as expected given the large proportion of variance explained by these variables. Adding additional effects resulted in only minimal changes in prediction accuracy (*R*^2^ = 0.211 for Model 2, *R*^2^ = 0.213 for Model 3, *R*^2^ = 0.218 for Model 4, and *R*^2^ = 0.221 for Model 5 and Model 6).

## DISCUSSION

In this study, we sought to decompose the phenotypic variance for three BP traits (DP, SP, PP) into components attributed to demographics, genetics, lifestyle and gene-by-lifestyle interactions, using a multi-ancestry subset of the UK Biobank. A multi-ancestry setting is prone to bias in variance component estimates due to population structure and sample imbalance. We addressed this challenge by constructing a structure-adjusted genomic relationship matrix using PC-Relate^31^, ensuring balanced sample sizes across major ancestry groups, and including the top genetic principal components in the statistical models. Our results showed that the additive genetic effect explains 10-18 % of the phenotypic variance for the BP traits. These estimates are a little lower than previous estimates of the genomic heritability based on individuals of European descent for these traits 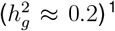^1^. However, this observation may be due to the difference in the number of genetic variants used – approximately 300,000 in our study and approximately 7,000,000 in the previous study. We also showed that lifestyle contributes moderately (∼10%) to DP variance, modestly (∼5%) to SP variance, and negligibly (∼1%) to PP variance. These findings are in agreement with recent studies using individuals of European descent^37^.

Our study showed that gene-by-lifestyle interactions explained ∼7%, ∼4%, and ∼3% of variance in DP, SP, and PP, respectively. Importantly, our estimates of the proportion of BP variance explained by G × E are much larger than those obtained in a recent study that used similar methodology and environmental variables to the present study, but only individuals of European descent^37^. These results highlight that gene-environment interactions can explain a larger proportion of variance in multi-ancestry samples compared to single-ancestry ones, presumably due to the increased genetic and environmental heterogeneity in this setting^29^. For instance, allelic heterogeneity and ancestry-specific LD patterns can increase the detectability of context-dependent genetic effects, particularly when interactions involve variants with variable tagging efficiency across populations^20,26^. Similarly, lifestyle such as diet, physical activity, and socioeconomic conditions may vary systematically by ancestry (as partly shown in Figure 3), making it easier to capture interactions in a multi-ancestry setting^9^. It is important to note that the estimates of the variance components other than the interaction one remained stable between Models 5 and 6. Thus, when included in Model 6, gene-by-lifestyle interactions explained variance that was instead unexplained in Model 5, rather than eroding variance from other components. Taken together, these contributions validate the utility of G × E models for capturing context-dependent genetic effects that are otherwise unmodeled in additive frameworks.

To further dissect the genetic architecture of G × E, we performed genome-wide interaction scans for each lifestyle variable. While no trait–lifestyle variable combination yielded a large number of genome-wide significant hits under a strict Bonferroni correction, we applied an elbow-point-based thresholding strategy to QQ plots, enabling the prioritization of interactions between genetic variants and lifestyle variables that exhibited signs of potential true association. The variants involved in the interactions revealed patterns of blood metabolites and vascular biology. For example, PKN2, a protein kinase involved in vascular tone regulation, is associated with systolic pressure^69^. PKN2-AS1, an antisense RNA to PKN2, is associated with red blood cell counts and sphingomyelin lipid levels^57,72^. Gene-level analysis revealed further biological connections to immune system function and neurological disorders, while providing additional support for blood metabolites and vascular biology. For example, TLR10, a Toll-like receptor, is responsible for downregulating immune responses in mammals^89^. TLR10 has known associations to other TLR protein levels, asthma, and peripheral artery disease^62,63,90^. Additionally, AP2A2 of the Adaptor complex is associated with cadherin levels, asthma, Alzheimer’s Disease and nicotine withdrawal complications^64–66,91^. Cadherins are calcium adhesion based molecules that have associations with blood pressure interactions with physical activity and vascular remodeling^92,93^. GO enrichment analyses supported these biological links, identifying T cell proliferation, Golgi apparatus packaging, and voltage-gated ion channels. Each of these functions has established connections to blood pressure or hypertension^89,94,95^. Thus, the individual gene-by-lifestyle interactions underlying the G × E variance exhibit plausible biological connections to blood pressure. However, given that our sample size is highly underpowered to reliably detect interactions, these results should be taken with great care. Additional work with larger sample sizes is needed to confirm our findings.

While G × E explained a substantial proportion of variance, its inclusion in the statistical model did not improve out of sample predictive accuracy. Across both random and cross-ancestry cross-validation schemes, we observed consistent trends: prediction performance was highest when including both genetic and lifestyle effects (Model 5) for DP and SP, underscoring the importance of combining genetic and lifestyle main effects. For PP, there was much less difference in accuracy among the six models, mainly due to demographic variables explaining much more variance than the other effects and driving the predictions. However, extending the model to include genome-wide G × E interactions (Model 6) did not result in further predictive gains. This plateau was observed across all three traits—DP, SP, and PP—and in both cross-validation scenarios.

This discrepancy between variance explained and predictive performance is likely due to several factors. First, the sample size in our study is very small, which limits the accuracy with which interaction effects to be used for prediction are estimated. Previous studies suggested that detecting robust G × E signals and translating them into predictive gain requires a much larger sample size^96,97^. Second, while the RKHS approach used in this study allows to include interaction effects easily, it assumes that the interactions between all genetic variants and all lifestyle variables affect the trait analyzed. This assumption very likely does not match the actual architecture of complex traits, resulting in the inclusion of a lot of noise in the model and, hence, poor prediction accuracy. Third, our models employed linear kernels, which may fail to capture nonlinear or threshold-based G × E effects^98^. Thus, it is possible that with larger sample sizes and more sophisticated statistical models gene-environment interactions may help improve cross-ancestry prediction accuracy^23^. More work is needed to confirm this hypothesis.

In conclusion, our findings provide evidence that G × E contribute more variance to BP traits in a multi-ancestry cohort compared to using only individuals of European descent. These results highlight the importance of modeling G × E in diverse populations, both to refine our understanding of trait architecture and to advance equitable precision medicine. In this light, efforts to assemble large and ethnically diverse data sets will be fundamental to understand the genetic architecture of complex human traits more thoroughly.

## Data Availability

The data used in our analyses are available from UK Biobank (https://www.ukbiobank.ac.uk).

## RESOURCE AVAILABILITY

### Lead contact

Requests for further information and resources should be directed to and will be fulfilled by the lead contact, Fabio Morgante.

### Materials availability

This study did not generate new materials.

### Data and code availability

The genotype and phenotype data used in our analyses are available from UK Biobank (https://www.ukbiobank.ac.uk/). Code used in the analyses is available at https://github.com/morgantelab/GxE_multi_ancestry_humans.

## ACKNOWLEDGMENTS

This research was conducted using the UK Biobank Resource under application number 62347. Research reported in this publication was supported by the National Institute of General Medical Sciences of the National Institutes of Health under Award Number R35GM146868 to FM. The content is solely the responsibility of the authors and does not necessarily represent the official views of the National Institutes of Health.

## AUTHOR CONTRIBUTIONS

Conceptualization: F.M.; Data curation: K.G., F.T., F.M.; Formal analysis: K.G., N.K.A.; Funding acquisition: F.M.; Investigation: K.G., N.K.A., F.T., T.F.C.M., F.M.; Methodology: K.G., F.T., T.F.C.M., F.M.; Project administration: F.M.; Resources: F.M.; Software: K.G., N.K.A., F.T.; Supervision: F.T., T.F.C.M., F.M.; Validation: K.G., N.K.A., F.T., T.F.C.M., F.M.; Visualization: K.G., N.K.A.; Writing-–original draft: K.G., N.K.A., F.M.; Writing-–review & editing: K.G., N.K.A., F.T., T.F.C.M., F.M.

## DECLARATION OF GENERATIVE AI AND AI-ASSISTED TECHNOLOGIES

During the preparation of this work, the author(s) used ChatGPT and Scite to source the literature and plan the flow of the manuscript. After using this tool or service, the authors reviewed and edited the content as needed and take full responsibility for the content of the publication.

