## Supplementary table and figures for "Gene-environment interactions contribute to blood pressure variation across global populations"

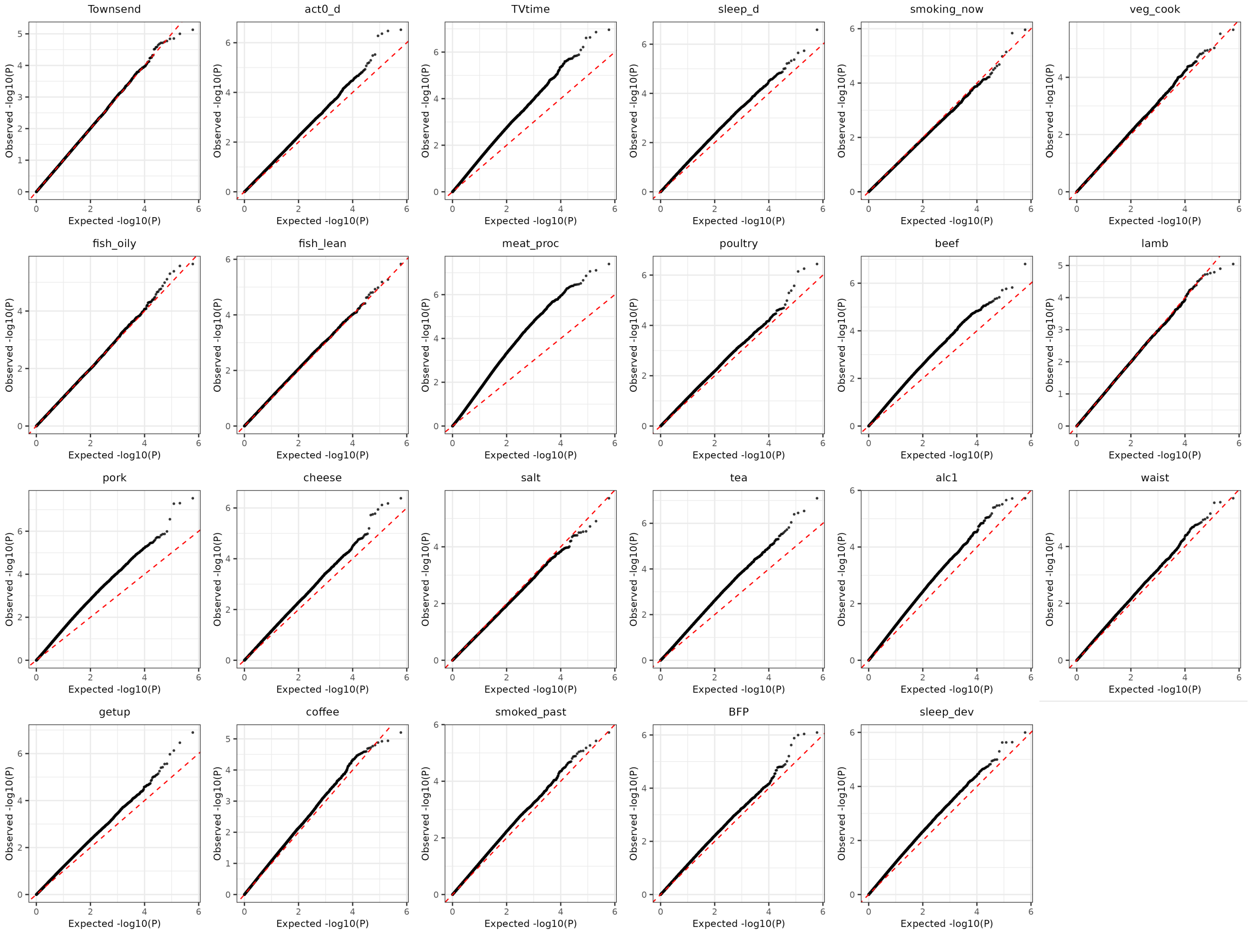


Figure S 1: QQ plots for G×E-GWAS for diastolic pressure.

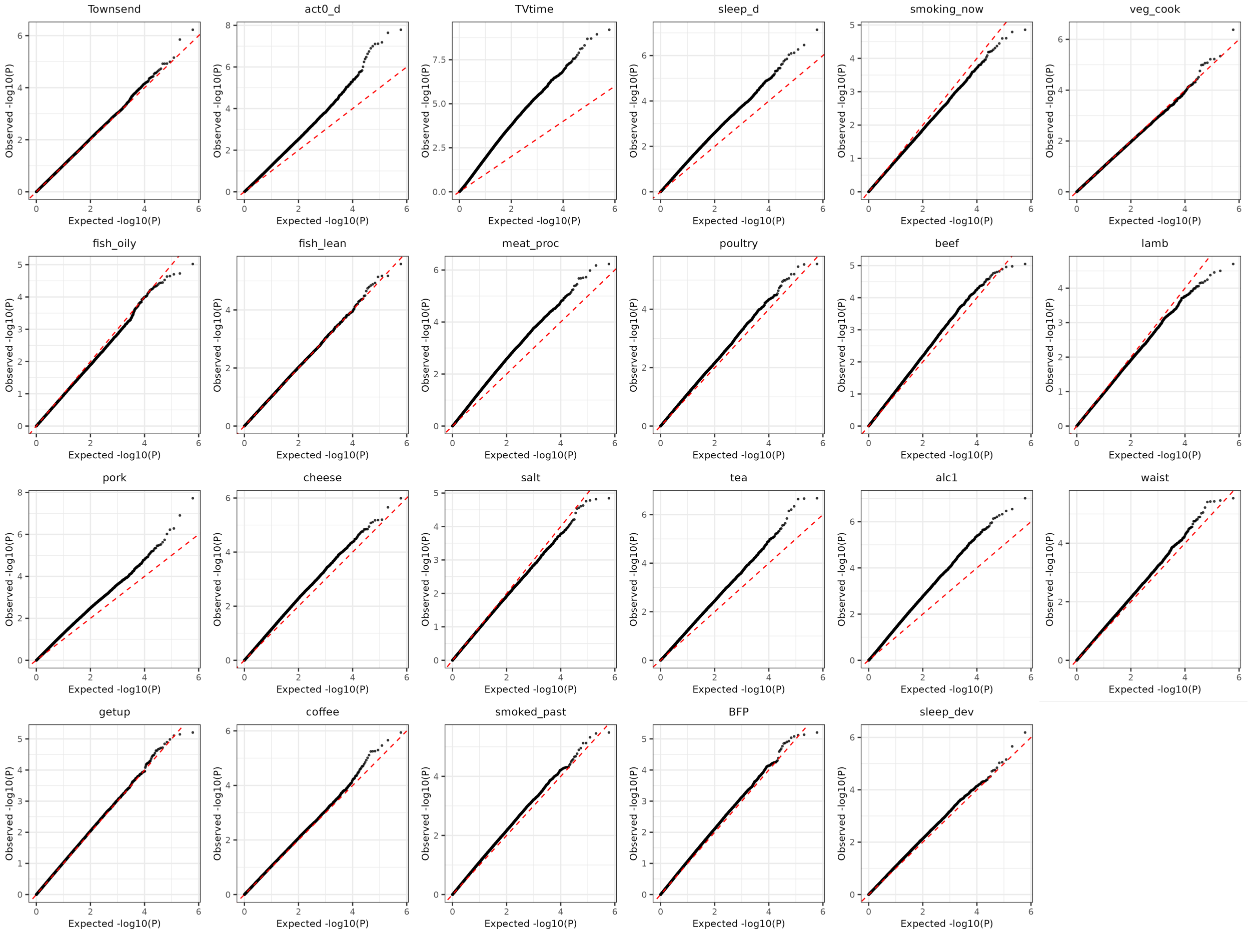


Figure S 2: QQ plots for G×E-GWAS for systolic pressure.

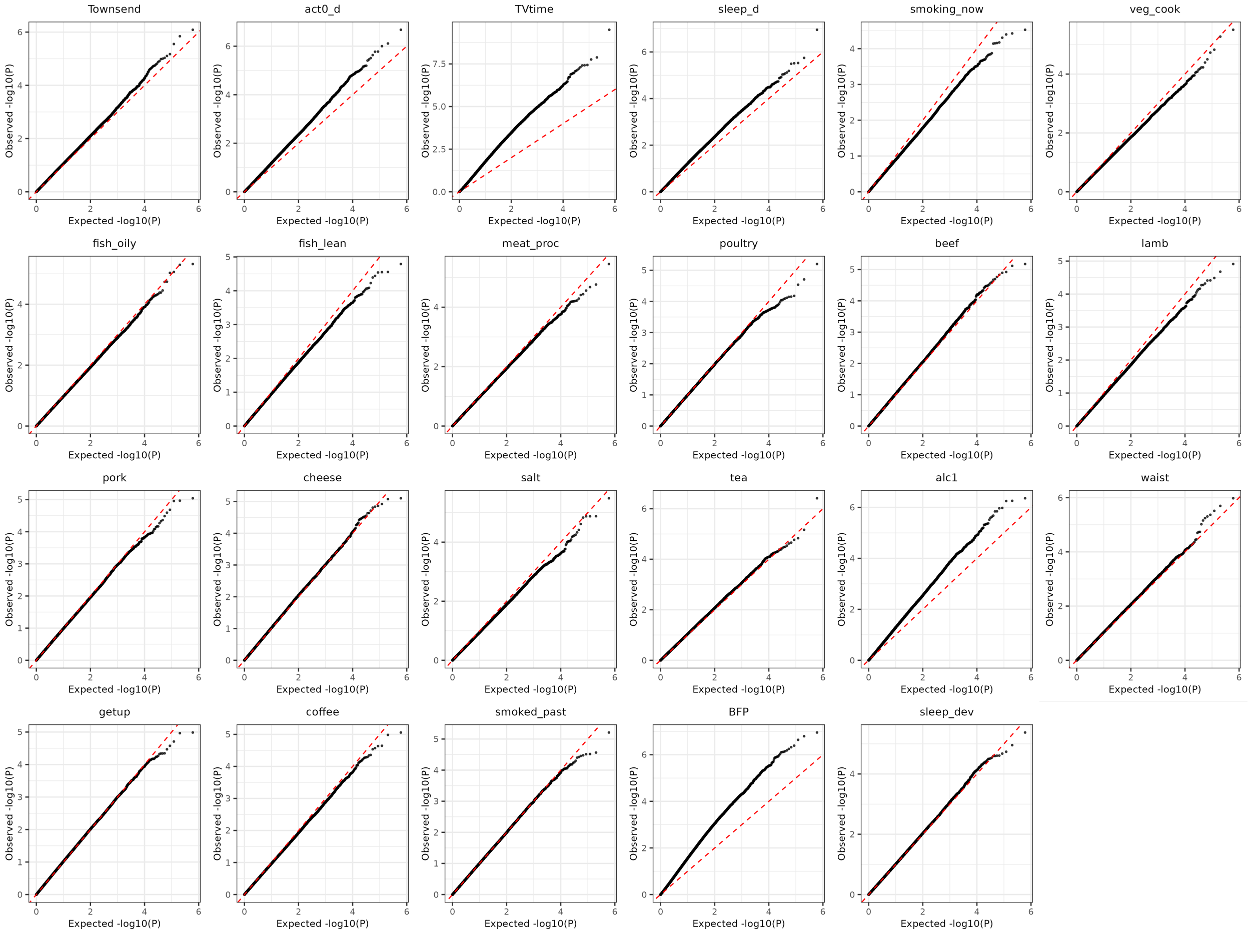


Figure S 3: QQ plots for G×E-GWAS for pulse pressure.

| Trait | Lifestyle | Variant ID | p-value |
|---|---|---|---|
| DP | act0_d | rs11246363 | 6.78985E-06 |
| DP | act0_d | rs12061410 | 6.28963E-06 |
| DP | act0_d | rs17567 | 3.3603E-07 |
| DP | act0_d | rs35086974 | 4.33246E-07 |
| DP | act0_d | rs4675569 | 5.11974E-06 |
| DP | act0_d | rs59930455 | 2.99218E-06 |
| DP | act0_d | rs60515720 | 3.28804E-06 |
| DP | act0_d | rs7308 | 5.29418E-07 |
| DP | act0_d | rs79030744 | 3.00886E-07 |
| DP | act0_d | rs7952081 | 8.52507E-06 |
| DP | BFP | rs1003167 | 4.26429E-05 |
| DP | BFP | rs11121855 | 1.62922E-05 |
| DP | BFP | rs12614105 | 1.58514E-05 |
| DP | BFP | rs1474813 | 0.000027963 |
| DP | BFP | rs1699670 | 2.10971E-05 |
| DP | BFP | rs17190392 | 2.14674E-05 |
| DP | BFP | rs2761413 | 0.000012689 |
| DP | BFP | rs282290 | 4.04483E-05 |
| DP | BFP | rs34917906 | 8.06632E-07 |
| DP | BFP | rs4863953 | 1.56702E-05 |
| DP | BFP | rs556852 | 1.00523E-06 |
| DP | BFP | rs57606675 | 1.58324E-05 |
| DP | BFP | rs60141239 | 2.88944E-05 |
| DP | BFP | rs62398131 | 1.39852E-05 |
| DP | BFP | rs6547747 | 4.28285E-05 |
| DP | BFP | rs72858315 | 0.000015744 |
| DP | BFP | rs74656059 | 1.67197E-05 |
| DP | BFP | rs7686574 | 6.24199E-06 |
| DP | BFP | rs77502768 | 9.15222E-07 |
| DP | BFP | rs7911984 | 1.31506E-06 |
| DP | BFP | rs8052204 | 0.000035761 |
| DP | BFP | rs9344830 | 3.76647E-05 |
| DP | BFP | rs9344849 | 2.39104E-06 |
| DP | BFP | rs978520 | 9.7075E-06 |
| DP | cheese | rs34334163 | 6.38121E-06 |
| DP | cheese | rs3788146 | 1.78686E-06 |
| DP | cheese | rs4485401 | 1.14667E-06 |
| DP | cheese | rs4566193 | 1.67643E-06 |
| DP | cheese | rs4781668 | 4.14463E-07 |
| DP | cheese | rs56122646 | 1.89959E-06 |
| DP | cheese | rs6702527 | 7.62207E-07 |
| DP | cheese | rs7555700 | 6.62523E-07 |
| DP | pork | rs4712266 | 4.96488E-08 |
| DP | pork | rs60032227 | 2.76692E-07 |
| DP | pork | rs6719772 | 5.25168E-08 |
| DP | pork | rs7192164 | 1.02269E-06 |
| DP | pork | rs8029848 | 2.93231E-08 |
| DP | poultry | rs10844998 | 3.62903E-07 |
| DP | poultry | rs11651728 | 1.03492E-05 |
| DP | poultry | rs1419028 | 7.17056E-07 |

| | | | |
|---|---|---|---|
| DP | poultry | rs1797517 | 5.10741E-06 |
| DP | poultry | rs28550256 | 5.55753E-07 |
| DP | poultry | rs55738317 | 2.66894E-06 |
| DP | poultry | rs80274100 | 1.49158E-05 |
| DP | poultry | rs9830867 | 4.15173E-06 |
| PP | sleep_d | rs10227705 | 1.11511E-07 |
| PP | sleep_d | rs1412948 | 2.97737E-06 |
| PP | sleep_d | rs241304 | 1.79939E-06 |
| PP | sleep_d | rs55941901 | 3.21104E-06 |
| PP | sleep_d | rs62177217 | 3.06425E-06 |
| PP | Townsend | rs17094899 | 6.72365E-06 |
| PP | Townsend | rs36107966 | 1.42865E-06 |
| PP | Townsend | rs4282989 | 2.80559E-06 |
| PP | Townsend | rs4532075 | 7.88658E-06 |
| PP | Townsend | rs79135558 | 8.16553E-07 |
| PP | waist | rs10940637 | 0.000019912 |
| PP | waist | rs10954734 | 1.05226E-06 |
| PP | waist | rs1213651 | 1.83008E-05 |
| PP | waist | rs1431140 | 4.94183E-06 |
| PP | waist | rs1866947 | 9.57283E-06 |
| PP | waist | rs1949207 | 1.85093E-05 |
| PP | waist | rs4356947 | 2.01229E-06 |
| PP | waist | rs4569287 | 5.70283E-06 |
| PP | waist | rs6467965 | 3.06641E-06 |
| PP | waist | rs6662147 | 6.96079E-06 |
| PP | waist | rs7229059 | 4.29257E-06 |
| SP | act0_d | rs10946345 | 5.7093E-07 |
| SP | act0_d | rs11246363 | 2.28095E-08 |
| SP | act0_d | rs12683372 | 6.5501E-08 |
| SP | act0_d | rs5007171 | 9.79593E-07 |
| SP | act0_d | rs55991456 | 2.34207E-07 |
| SP | act0_d | rs609602 | 3.35419E-07 |
| SP | act0_d | rs72684239 | 1.62832E-08 |
| SP | act0_d | rs72943699 | 1.80208E-07 |
| SP | act0_d | rs73171619 | 1.00547E-07 |
| SP | act0_d | rs78301771 | 7.65523E-08 |
| SP | act0_d | rs79030744 | 1.26978E-07 |
| SP | act0_d | rs7942693 | 7.85334E-08 |
| SP | act0_d | rs9521815 | 4.13927E-07 |
| SP | coffee | rs1030130 | 3.03031E-05 |
| SP | coffee | rs10928858 | 2.48915E-05 |
| SP | coffee | rs11964975 | 1.18778E-05 |
| SP | coffee | rs12418914 | 4.46511E-05 |
| SP | coffee | rs12666634 | 5.67766E-06 |
| SP | coffee | rs13015031 | 1.67132E-05 |
| SP | coffee | rs13053817 | 1.14698E-06 |
| SP | coffee | rs1642822 | 1.94395E-05 |
| SP | coffee | rs16987627 | 2.20096E-06 |
| SP | coffee | rs1791411 | 4.36633E-05 |
| SP | coffee | rs2027667 | 0.000033523 |
| SP | coffee | rs3817648 | 3.44061E-06 |

| | | | |
|---|---|---|---|
| SP | coffee | rs3847015 | 2.22683E-05 |
| SP | coffee | rs4709392 | 3.88719E-05 |
| SP | coffee | rs4716650 | 9.71743E-06 |
| SP | coffee | rs4717878 | 7.72502E-06 |
| SP | coffee | rs57740470 | 5.52886E-06 |
| SP | coffee | rs60423784 | 3.40392E-05 |
| SP | coffee | rs62095715 | 1.43394E-05 |
| SP | coffee | rs72844231 | 5.04221E-06 |
| SP | coffee | rs7567596 | 5.60283E-06 |
| SP | coffee | rs77356523 | 2.60573E-05 |
| SP | coffee | rs77356619 | 4.36954E-05 |
| SP | coffee | rs8104843 | 1.98363E-05 |
| SP | coffee | rs9346670 | 4.74978E-05 |
| SP | tea | rs10166913 | 6.23479E-07 |
| SP | tea | rs10795917 | 4.55289E-07 |
| SP | tea | rs12314904 | 1.44187E-06 |
| SP | tea | rs16972467 | 2.11729E-07 |
| SP | tea | rs41345745 | 2.29668E-07 |
| SP | tea | rs41411848 | 2.18277E-07 |
| SP | tea | rs7079622 | 7.07788E-07 |
| SP | Townsend | rs17094899 | 0.000001412 |
| SP | Townsend | rs3786180 | 5.94476E-07 |
| SP | veg_cook | rs10234443 | 6.03943E-06 |
| SP | veg_cook | rs10238894 | 8.5972E-06 |
| SP | veg_cook | rs12145922 | 5.96863E-06 |
| SP | veg_cook | rs2079868 | 4.58437E-06 |
| SP | veg_cook | rs4520040 | 1.02595E-05 |
| SP | veg_cook | rs6480394 | 8.32301E-06 |
| SP | veg_cook | rs7137797 | 1.00074E-05 |
| SP | veg_cook | rs786906 | 0.000017527 |
| SP | veg_cook | rs795443 | 4.18308E-07 |

Table S 1: List of genetic variants and lifestyle variables selected using the QQ plots from G×E-GWAS for each blood pressure trait.

| Trait | Lifestyle | Variant ID | Gene | Description | Citation |
|---|---|---|---|---|---|
| DP | act0_d | rs4675569 | PPIAP68, RN7SKP178 | Striated muscle fiber area | [1] |
| DP | act0_d | rs59930455 | FOXO3, LINC00222 | mosquito bite reaction | [2] |
| DP | BFP | rs11121855 | TNFRSF8 | Gut microbe interactions | [3] |
| DP | cheese | rs4485401 | CNTNAP4 | Sphingolipid levels | [4] |
| DP | cheese | rs4781668 | BMERB1 | phosphatidylcholine levels | [5] |
| DP | cheese | rs4781668 | BMERB1 | phosphatidylcholine levels | [5] |
| DP | cheese | rs4781668 | BMERB1 | cholesterol ester levels | [5] |
| DP | cheese | rs4781668 | BMERB1 | height | [6] |
| DP | pork | rs4712266 | ARPC3P5, LINC02543 | Femoral neck bone mineral density | [7] |
| DP | pork | rs6719772 | LRRTM4, RN7SKP164 | lipid levels in elite athletes | [8] |
| DP | poultry | rs1419028 | SPOCK3-x-RPSAP72, TENT5A | PHF-Tau interaction | [9] |
| SP | coffee | rs13053817 | RFPL1S | carotid artery thickness, hiv infection sclerosis | [10] |
| SP | coffee | rs13053817 | RFPL1S, RFLP1, NEFH | acute myeloid leukemia core binding factor | [11] |
| SP | coffee | rs4709392 | IGF2R | CPVL protein levels in blood | [12] |
| SP | tea | rs10795917 | UPF2-x-SPINT2 | prostate cancer snps x snp interaction | [13] |
| SP | veg_cook | rs12145922 | PKN2-AS1 | liver enzyme gamma glutamyl transferase levels | [14] |
| SP | veg_cook | rs12145922 | PKN2-AS1 | RBC count erythrocyte | [15] |
| SP | veg_cook | rs12145922 | PKN2-AS1 | RBC count erythrocyte | [15] |
| SP | veg_cook | rs12145922 | PKN2-AS1 | sphingomyelin levels in elite athletes | [8] |
| SP | veg_cook | rs4520040 | R3HDM2P2, GRIK2 | cardiac hypertrophy | [16] |
| SP | veg_cook | rs786906 | PKN2 | Systolic blood pressure | [17] |
| SP | veg_cook | rs786906 | PKN2 | Intraocular pressure | [18] |

Table S 2: List of selected variants with known associations in the GWAS Catalog.

| Trait | Gene | Description | DP | SP | PP | CV | BM | IS | ND |
|---|---|---|---|---|---|---|---|---|---|
| DP | ABLIM2 | AFAP1 protein levels (actin filament assoc.) | | | | 12 | | | |
| DP | AC104695.4 | | | | | | | | |
| DP | ACOT7 | several blood metabolites (KYAT1, PLPBP, QDPR, FKBP4), waist | | | | | 19 | | |
| DP | AKR1D1 | bone density, testosterone | | | | | | | |
| DP | AP2A2 | cadherin levels, alzheimers, nicotine withdrawal | | | | | 12 | 20 | 21 |
| DP | ARPC3P5 | blood metabolites, bone density, heart rate | | | | 22 | 23 | | |
| DP | C16orf45 | | | | | | | | |
| DP | CLEC2B | CLEC7A levels, bone density, LDL in schizophrenia | | | | | 12 | | 24 |
| DP | CNTNAP4 | Smoking, intelligence, cholesterol | | | | | 25 | | 26 |
| DP | CREB3L2 | Height, cholesterol, pulse pressure | | | 27 | | 28 | | |
| DP | EBF3 | restless leg syndrome | | | | | | | 29 |
| DP | EPS15 | Atrial fibrillation, stroke, pulse pressure | | | 30 | 31 | | | |
| DP | FFAR4 | Height, vitamin A, triglycerides | | | | | 32 | | |
| DP | IPO5 | liver proteins, gut microbiome, sleep duration | | | | | 12 | | 33 |
| DP | ITGAE | ITGAV/ITGB7 protein level ratio, Parkinsons, ADHD | | | | | 19 | | 34 |
| DP | ITGB2 | several protein ratios (ITGB1, ADGRE5, ITGAM, IGF1R), cholesterol | | | | | 19 | | |
| DP | KCNMB3 | Corpuscular hemoglobin, antidepressant response, trypanosome EKG PR | | | | 35 | | | 36 |
| DP | KLRF2 | CLEC7A protein levels, immune protein levels, LDL in schizophrenia | | | | | | 37 | 24 |
| DP | KRCC1 | | | | | | | | |
| DP | MS4A18 | TREM2 protein levels, gut microbiome | | | | | 12 | | |
| DP | NAALADL2 | height, education, smoking, pulse pressure | | | 38 | | | | 26 |
| DP | PLB1 | self levels, eosionophils, heart rate, systolic pressure | | 15 | | 15 | 12 | 15 | |
| DP | PNP | spinal fluid protein levels, Alzheimer's | | | | | | 39 | |
| DP | RAB19 | height, kidney function, cheese | | | | | | | |
| DP | RN7SKP40 | | | | | | | | |

| | | | | | | | | | |
|---|---|---|---|---|---|---|---|---|---|
| DP | RNGTT | height, AML, sleep apnea, BPD, depression | | | | | 12 | | 40 |
| DP | RNU6-1281P | | | | | | | | |
| DP | RORA | 500+ hits: SP, PP, intraocular pressure, liver enzyme levels | | 41 | 30 | 15 | 42 | 43 | 44 |
| DP | RP11-191G24.1 | | | | | | | | |
| DP | RP11-290C10.1 | | | | | | | | |
| DP | RP11-365F18.3 | | | | | | | | |
| DP | RP11-775H9.2 | | | | | | | | |
| DP | SLC30A8 | T2D, hemoglobin, intraocular pressure, several neuro disorders (scz, self-harm) | | | | | 45 | | 46 |
| DP | TENM3 | bone density, AML, smoking, neuroticism, neuro disorders, varicose veins, SP | | 47 | | 48 | | 49 | 50 |
| DP | TLR10 | tlr levels, asthma, peripheral arterial disease | | | | 51 | | 52 | |
| DP | TMEM51 | Migraine, DP, SP, PP | 30 | 30 | 30 | | | | 53 |
| DP | TMEM51-AS1 | Glomelular function, postural challenges, neurofibrillary tangles (Tau protein), CCL20 | | | | | | 54 | 9 |
| DP | TMEM55B | | | | | | | | |
| DP | TNFRSF8 | Self, eosinophil levels, asthma | | | | | 12 | 55 | |
| DP | TTC26 | | | | | | | | |
| DP | VSTM4 | brain groove depth, platelet count, migraine | | | | 56 | | | 57 |
| DP | WNT7B | connective tissue disease, Hematocrit, vision loss, myopia, male | | | | 56 | | | |
| DP | ZDHHC14 | platelet characteristics (size, volume, maturity), smoking ptsd, ADHD, MDD | | | | 58 | 19 | | 59 |
| PP | CACNA2D1 | growth hormone levels, maternal schizophrenia, SP, BPD | | 38 | | | | | 15 |
| PP | CLMP | hemoglobin, heart rate, RBC, SP, PP, DP | 38 | 30 | 30 | 15 | 19 | | |
| PP | CTD-2216M2.1 | | | | | | | | |
| PP | CTD-2332E11.2 | | | | | | | | |

| | | | | | | | | | |
|---|---|---|---|---|---|---|---|---|---|
| PP | DAB1 | tumor calcium signalling, restless leg, education, smoking, anti hypertensives, blood proteins, insomnia | | | | 60 | 12 | 15 | 29 |
| PP | LRRC8D | platelet, cognitive, lung cancer, depression, OCD, gut microbio | | | | 56 | 12 | | 61 |
| PP | MIR4780 | | | | | | | | |
| PP | OSBPL6 | smoking, EKG, SP/DP, alzheimers, myopia | 62 | 62 | | 63 | | | 64 |
| PP | RP11-91I11.1 | | | | | | | | |
| PP | SMYD1 | bone, alanine levels, night sleep phenotypes, neurofibrillary tangles, retinal detachment (eye), VEGFA | | | | 54 | | | 65 |
| SP | AFTPH | height, cognition, microbiomes | | | | | | | |
| SP | AP2A2 | cadherin levels, alzheimers, nicotine withdrawal | | | | | 12 | 20 | 21 |
| SP | C2orf71 | | | | | | | | |
| SP | CD6 | self/cd5 levels, autoimmune diseases, T1D, | | | | | 19 | 19 | |
| SP | CDCP1 | immune protein levels, eye degeneration, alzheimers | | | | | 19 | 66 | |
| SP | COL4A2 | Several types of heart disease, DP, PP | 41 | | 67 | 68 | | | |
| SP | CTIF | height, hematocrit, heart disease, smoking, autoimmune, DP/SP | 62 | 62 | | 56 | | 56 | |
| SP | ELMO1 | platelet characteristics, eosinophils/lympocytes, several autoimmune, smoking, ADHD, BMI, MS | | | | 12 | | 56 | 69 |
| SP | GADL1 | beta alanine and N-acetylcarnosine levels, AML, glomelular function | | | | | 12 | | |
| SP | HK1 | hemolgobin, hematocrit, cholesterol | | | | 58 | 45 | | |
| SP | IGF2R | blood protein levels, lipid levels, CAD, | | | | 70 | 19 | | |
| SP | MUC2 | lung disease, asthma, cholesterol/lipids, eosinophil, alcohol | | | | | 12 | 71 | |
| SP | NCAPD2 | blood protein levels, bone density | | | | | 72 | | |
| SP | OR1I1 | PGLYRP2 protein levels, DP/SP, gut microbio, smoking, carotid thickness | 62 | 62 | | 73 | 12 | | |
| SP | PAMR1 | self(muscle regen), height, longevity down | | | | | 12 | | |

| | | | | | | | | | |
|---|---|---|---|---|---|---|---|---|---|
| SP | PKN2 | immune system GBP4, height, SP, PP, DP | 74 | 15 | 45 | | 12 | 12 | |
| SP | PLA2G4E | neutrophils, platelet volume, smoking | | | | 12 | | 56 | |
| SP | PPM1H | RBC characteristics, basinophil counts, microbiome, OCD, heart failure, height/bone | | | | 56 | | 15 | 75 |
| SP | RFPL1 | Acute Myeloid Leukemia | | | | | | | |
| SP | RFPL1S | bone density, blood characteristics, cognitive, T2D, eye disease | | | | 76 | 12 | | |
| SP | RNF165 | | | | | | | | |
| SP | RNU6-933P | | | | | | | | |
| SP | RP11-187O7.3 | | | | | | | | |
| SP | RP11-227H15.7 | | | | | | | | |
| SP | RP11-503C24.2 | | | | | | | | |
| SP | RP11-503C24.3 | | | | | | | | |
| SP | RP11-76N22.2 | | | | | | | | |
| SP | SCARB1 | 500+ HDL levels, coronary artery disease | | | | 12 | 77 | | |
| SP | SCARNA10 | | | | | | | | |
| SP | TAF3 | bone, magnesium, RBC, neurofibrillary tangles, smoke, peripheral artery disease, hearing loss | | | | 56 | | 9 | |
| SP | TLR10 | tlr levels, asthma, peripheral arterial disease | | | | 51 | | 52 | |
| SP | TXNDC16 | bromotryptophan levels, tinnitus, Alzheimer's, arthritis, sleep loss, memory loss, | | | | | 78 | 79 | 80 |
| SP | UPF2 | height, liver proteins, gut microbio, gut wall diverticulitis, | | | | | 12 | | |

Table S 3: List of genes identified by Variant Effect Predictor (VEP) for each blood pressure trait. Descriptions summarize the top GWAS Catalog associations for the respective gene. References for the known associations with diastolic pressure (DP), systolic pressure (SP), pulse pressure (PP), cardiovascular system (CV), blood metabolites (BM), immune system (IS), and neurological disorders (ND) are provided in the relevant column.

| Trait | Source | Term | ID | p-value | Intersections |
|---|---|---|---|---|---|
| DP | GO:CC | plasma membrane | GO:0005886 | 0.041469468 | CLEC2B, CNTNAP4, EPS15, FFAR4, ITGAE, ITGB2, KCNMB3, KLRF2, MS4A18, RAB19, TENM3, TLR10, VSTM4, WNT7B, TNFRSF8, AP2A2, PLB1, SLC30A8 |
| DP | CORUM | PNP homotrimer complex | CORUM:6208 | 0.049679937 | PNP |
| SP | GO:CC | trans-Golgi network transport vesicle | GO:0030140 | 0.002226064 | AFTPH, AP2A2, IGF2R |
| SP | GO:CC | clathrin coat | GO:0030118 | 0.005027312 | AFTPH, AP2A2, IGF2R |
| SP | GO:CC | AP-1 adaptor complex | GO:0030121 | 0.018621239 | AFTPH, AP2A2 |
| SP | GO:CC | Golgi-associated vesicle | GO:0005798 | 0.033354012 | AFTPH, AP2A2, IGF2R |
| SP | GO:CC | membrane coat | GO:0030117 | 0.03653315 | AFTPH, AP2A2, IGF2R |
| SP | GO:CC | coated membrane | GO:0048475 | 0.03653315 | AFTPH, AP2A2, IGF2R |
| SP | GO:CC | clathrin coat of trans-Golgi network vesicle | GO:0030130 | 0.040654057 | AFTPH, AP2A2 |
| SP | GO:CC | trans-Golgi network transport vesicle membrane | GO:0012510 | 0.049856885 | AFTPH, AP2A2 |
| PP | GO:MF | voltage-gated calcium channel activity involved in bundle of His cell action potential | GO:0086057 | 0.049847069 | CACNA2D1 |
| PP | CORUM | LRRC8A-LRRC8D complex | CORUM:6582 | 0.049931693 | LRRC8D |

Table S 4: List of terms identified by Gene Ontology (GO) enrichment analysis for each blood pressure trait.
